# Interpretable machine learning for coeliac disease diagnosis: quantitative morphometry of duodenal biopsies

**DOI:** 10.64898/2026.06.02.26354731

**Authors:** Rebekah Bryant, Jacobo Romero Diaz, Adam G. Scott, Aryan A. Sagdeo, Gabriella Z. Jenkins, Robert A. Richardson, James Y. H. Chan, Mark J. Arends, Elizabeth J. Soilleux, Florian Jaeckle

## Abstract

**Background:** Coeliac disease affects approximately 1% of the global population and remains substantially underdiagnosed. Histopathological assessment of duodenal biopsies is the diagnostic gold standard but is subject to approximately 20% inter-observer disagreement. While machine learning approaches show promise, most prior work relies on black-box models with limited interpretability, restricting clinical adoption.

**Methods:** We present an interpretable pipeline that follows established histopathological criteria by extracting clinically meaningful morphological features from H&E-stained whole-slide images. Five sequential stages perform pre-processing, semantic segmentation of villi, crypts, intraepithelial lymphocytes (IELs) and enterocytes, crypt morphometry, villus length estimation via a novel polyline-based keypoint model, and coeliac disease classification using three quantitative features: IEL-to-enterocyte ratio, villus-to-crypt area ratio, and villus-length-to-crypt-depth ratio. Training and validation used data from four institutions; independent testing used 1,357 WSIs from two further institutions including one with a previously unseen scanner manufacturer, spanning five diagnostic categories: coeliac disease, normal mucosa, chronic inflammation, gastric metaplasia, and gastric heterotopia.

**Results:** Semantic segmentation achieved villus and crypt precision and recall of 87–90%. Villus length estimation correlated strongly with expert annotations (Pearson’s r=0.85, mean relative error 13.5% post-calibration). All three morphological features significantly separated coeliac disease from all non-coeliac diagnostic groups across internal and external datasets (p<0.01 in all comparisons). On the test set the diagnostic classifier achieved accuracy 94.5%, PPV 92.9%, NPV 94.7%, and AUC 0.982.

**Conclusions:** This interpretable framework achieves strong multi-centre diagnostic performance while producing quantitative morphological outputs, villus length, crypt depth, and IEL-to-enterocyte ratios, that directly reflect established histopathological criteria, representing a meaningful step towards standardised AI-assisted coeliac disease diagnosis.

## Introduction

Coeliac disease is a chronic autoimmune disorder triggered by the ingestion of gluten, a protein found in wheat, barley, and rye. It affects approximately 1% of the global population, although it remains substantially underdiagnosed [1]. Active coeliac disease is associated with a wide range of gastrointestinal and extraintestinal symptoms, including diarrhoea, abdominal pain, weight loss, anaemia, and fatigue [2]. In addition to its symptomatic burden, untreated disease increases the risk of serious complications, such as duodenal adenocarcinoma, lymphoma, and infertility [3]. Currently, the only effective treatment is lifelong adherence to a gluten-free diet, which can alleviate symptoms and reduce long-term risks [4].

The diagnosis of coeliac disease typically involves a combination of serological testing and histopathological assessment. Initial screening is performed using blood tests, most commonly tissue transglutaminase IgA (tTG-IgA). In cases where tTG-IgA levels are not at least ten-times the upper limit of normal, or when confirmatory testing is required, current clinical guidelines (e.g. NICE) recommend duodenal biopsy as the diagnostic gold standard [4]. Histopathological evaluation of haematoxylin and eosin (H&E)-stained biopsies focuses on key morphological features (Figure 2), including an increased number of intraepithelial lymphocytes (IELs) (immune cells located within the intestinal epithelium), villus atrophy (shortening of the finger-like projections lining the duodenum), and crypt hyperplasia (elongation of the glandular crypts at the base of the villi). An IEL count exceeding approximately 25–30 IELs per 100 enterocytes is considered indicative of coeliac disease, depending on the guideline used [5,6]. Similarly, a reduced villus-to-crypt length ratio reflects mucosal architectural distortion, with normal ratios of approximately 3:1–5:1 becoming markedly reduced in coeliac disease [7,8]. Accurate assessment of these features requires well-oriented biopsies, as the three-dimensional structure of the mucosa must be sectioned appropriately to allow reliable measurement [9].

**Figure 1.**
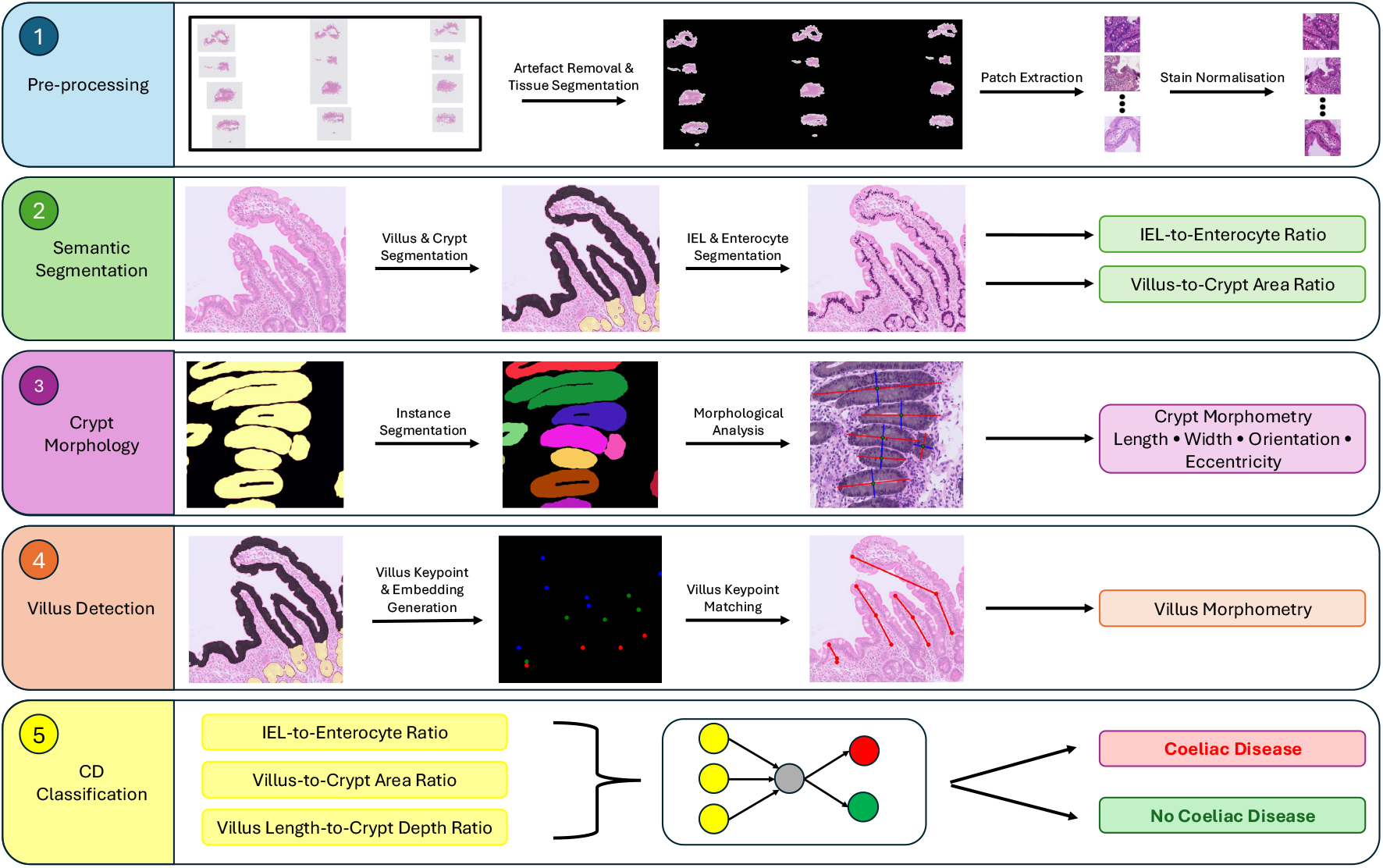
Overview of the proposed interpretable coeliac disease diagnosis pipeline. The pipeline consists of five sequential stages. (1) Pre-processing: WSIs undergo artefact removal and tissue segmentation, followed by patch extraction and stain normalisation. (2) Semantic segmentation: separate deep learning models segment villi and crypts, and IELs and enterocytes, enabling computation of IEL-to-enterocyte and villus-to-crypt area ratios. (3) Crypt morphology analysis: crypt semantic segmentation masks are converted into instance-level segmentations, from which morphological features including crypt length, width, orientation, and eccentricity are extracted. (4) Villus detection and morphology analysis: a polyline-based model predicts villus heatmaps and embedding representations, which are used to extract and match villus keypoints to form individual villus polylines and estimate villus morphology. (5) Coeliac disease classification: quantitative features derived from the previous stages, including IEL-to-enterocyte ratio, villus-to-crypt area ratio, and villus-length-to-crypt-depth ratio, are combined within a supervised classifier to predict coeliac disease status. WSI: whole slide image. IEL: intraepithelial lymphocyte.

**Figure 2.**
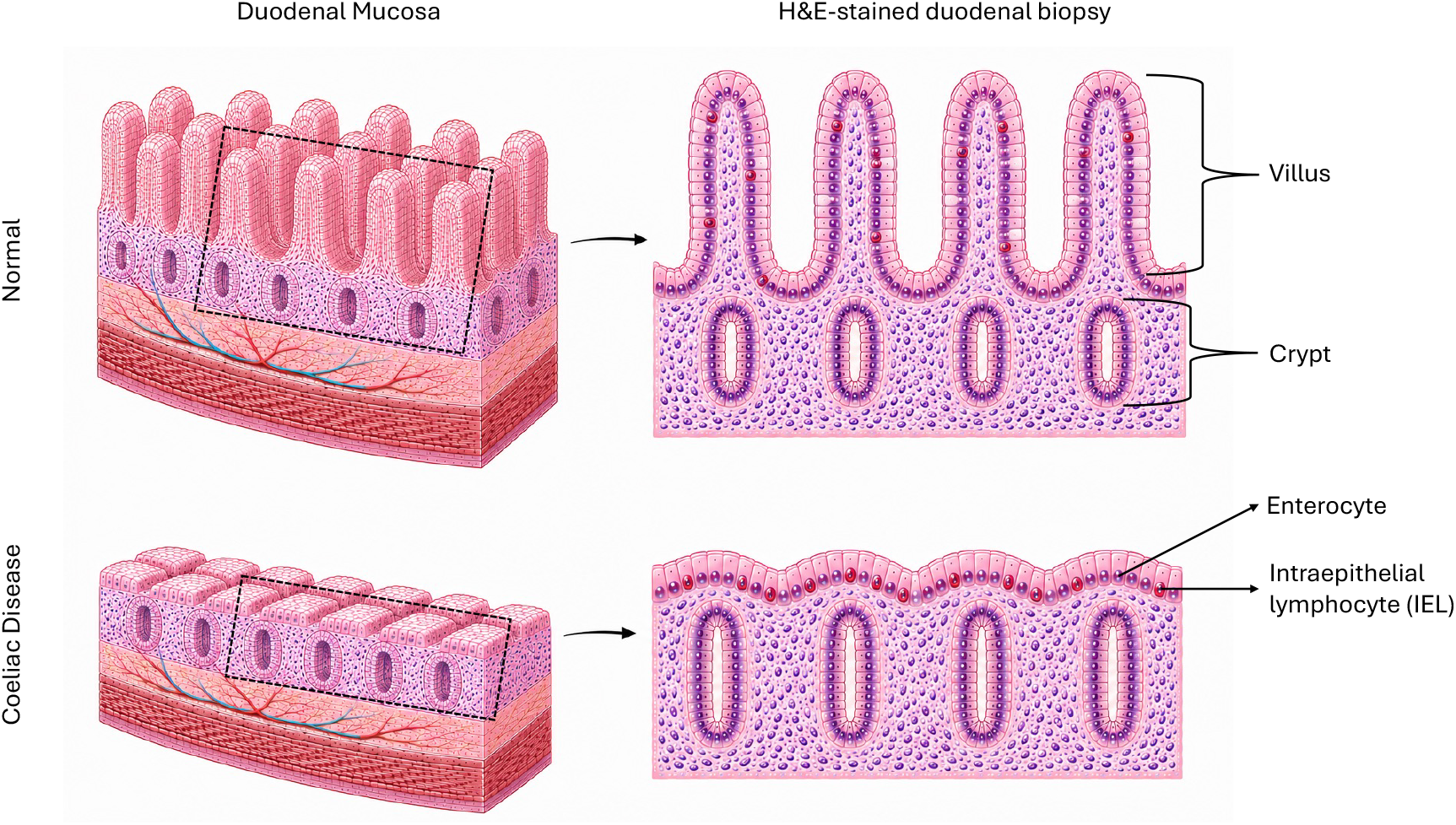
Schematic representation of the three-dimensional duodenal mucosa (left) and the cross-sectional haematoxylin and eosin (H&E)-stained duodenal biopsy appearance (right). The characteristic changes observed in coeliac disease include shortening of the villi (villous atrophy), lengthening of the crypts (crypt hyperplasia), and an increased number of intraepithelial lymphocytes (IELs) relative to the number of enterocytes.

Despite established diagnostic criteria, histopathological assessment of coeliac disease is subject to considerable inter-observer variability, with reported disagreement rates of approximately 20% among pathologists [10–15]. This variability highlights the need for objective, reproducible, and quantitative diagnostic tools.

In recent years, artificial intelligence (AI) approaches have been explored to assist in the diagnosis of coeliac disease. However, most prior work has relied on black-box models that offer limited interpretability [16–20]. Interpretability is increasingly recognised as important for clinical adoption of AI systems. In a JAMIA study, 88% of physicians preferred machine-learning outputs accompanied by explanations over unexplained predictions [21]. Similarly, 84.8% of US oncologists reported that AI models would need to be explainable for clinical use [22], while 56.3% of referring physicians in a radiology implementation survey identified model transparency as the most important factor for trust in AI systems [23]. These findings highlight the importance of developing AI systems that provide clinically meaningful and interpretable outputs rather than relying solely on black-box predictions.

A few papers have investigated diagnosing coeliac with interpretable AI. For example, Gruver *et al*. [24] employed a customised segmentation pipeline to identify villous epithelium, lamina propria, and crypts in H&E-stained biopsies, and IELs and enterocytes in CD3 immunohistochemistry whole-slide images (WSIs). While promising, this approach depends on CD3 staining, which is not routinely used in clinical practice, and was evaluated on data from a single source, limiting evidence of generalisability. Similarly, Griffin *et al*. [25] developed a convolutional neural network to identify tissue and cellular components and correlate extracted features with Marsh scores. However, their study has not been peer-reviewed and was limited by a small test cohort (n = 28) and the use of data from the same source for both training and evaluation. More recently, Tyagi et al. [26] proposed MeasureNet, a keypoint-based approach for measuring villus morphology using base–tip representations, achieving a reported classification accuracy of 82.66%. However, this work has not yet undergone peer review and did not evaluate generalisation across external institutions or scanner types.

In our previous work, we introduced a set of semantic segmentation models capable of detecting IELs, enterocytes, villi, and crypts in H&E-stained WSIs, and demonstrated generalisation to data from external sources [27]. Building on this foundation, the present study introduces several key advances.

First, we improve the accuracy of villus–crypt semantic segmentation by expanding the training dataset and adopting a more powerful backbone architecture. Second, we introduce a crypt morphological analysis framework that extracts instance-level features, including crypt length, width, orientation, and eccentricity, rather than relying solely on aggregate measurements. Third, we propose a polyline-based model for detecting individual villi and estimating their lengths via keypoint prediction and matching. Fourth, we develop an interpretable diagnostic classifier that integrates these quantitative features (IEL-to-enterocyte ratio, villus-to-crypt area ratio, villus length-to-crypt depth ratio) to distinguish coeliac disease from a range of non-coeliac diagnoses encountered in routine clinical practice, including normal mucosa, chronic inflammation, gastric metaplasia, and gastric heterotopia, and evaluate its performance on external datasets from previously unseen institutions and scanner types. Finally, we perform comprehensive ablation studies to evaluate the contribution of individual features and model components (Supplementary Material).

## Datasets

We used four distinct datasets for training, validation, and testing of our models. A summary of all datasets is provided in Table 1.

**Table 1.** The Villus–Crypt Semantic Segmentation dataset comprises 2048 × 2048 pixel patches at 40× magnification, whereas all other datasets consist of whole-slide images (WSIs). Case counts are stratified by diagnosis, source institution, scanner type, sex, and age. Demographic information (age and sex) was unavailable for a subset of cases, primarily from the North Tees and Addenbrooke’s cohorts.

| Tables |  | Villus-Crypt<br>Semantic<br>Segmen-tation<br>(n=94) | Villus<br>Polyline<br>(n=41) | Classifi-<br>cation<br>Validation<br>(n=545) | Classifi-<br>cation Test<br>(n=1357) |
| --- | --- | --- | --- | --- | --- |
| Diagnosis | Coeliac | 37 | 16 | 163 | 250 |
|  | Normal | 50 | 22 | 232 | 795 |
|  | Other | 7 | 3 | 150 | 312 |
| Source | Addenbrookes | 27 | 16 | 290 |  |
|  | North Tees | 21 | 8 | 80 |  |
|  | Warwick | 6 |  | 114 |  |
|  | Glasgow | 22 | 9 | 61 |  |
|  | Heartlands | 11 | 8 |  |  |
|  | Sheba | 7 |  |  |  |
|  | Edinburgh |  |  |  | 644 |
|  | Southampton |  |  |  | 713 |
| Scanner | Leica Aperio AT2 | 33 | 16 | 48 |  |
|  | Philips IntelliSite Ultra<br>Fast Scanner | 32 | 9 | 417 | 644 |
|  | Hamamatsu 210 | 21 | 8 | 80 |  |
|  | Roche Ventana iScan<br>HT | 5 | 8 |  |  |
|  | Hamamatsu<br>Nanozoomer XR | 3 |  |  |  |
|  | 3DHISTECH Kft.<br>Pannoramic 1000 |  |  |  | 713 |
| Sex | Female | 36 | 22 | 305 | 755 |
|  | Male | 25 | 10 | 240 | 582 |
| Age | Median | 42 | 55 | 60 | 49 |
|  | 0-9 | 0 |  | 5 | 123 |
|  | 10-19 | 0 |  | 25 | 315 |
|  | 20-29 | 10 | 5 | 53 | 55 |
|  | 30-39 | 15 | 4 | 62 | 66 |
|  | 40-49 | 13 | 6 | 65 | 127 |
|  | 50-59 | 4 | 4 | 63 | 173 |
|  | 60-69 | 8 | 6 | 103 | 218 |
|  | 70-79 | 7 | 3 | 111 | 182 |
|  | 80-89 | 4 | 4 | 54 | 95 |
|  | 90-99 | 0 |  | 6 | 3 |

### Semantic Segmentation Dataset

For the semantic segmentation of villi and crypts, we assembled a dataset comprising 94 image patches (2048 × 2048 pixels) at 40× magnification. All villi and crypt structures within these patches were manually annotated by one of four human annotators. The dataset included 51 normal cases, 36 CD cases, and 7 cases with other non-coeliac pathological diagnoses. Data were collected from six hospitals and digitized using five different scanners. Demographic information (age and sex) was available for 61 cases (65%), with a median age of 42 years and 59% female. Of the 94 patches, 54 were previously used in our earlier study [27]. We augmented this dataset with an additional 40 patches (23 normal, 17 CD, and 1 other diagnosis), manually annotated by a new annotator. These additional patches were selected with specific objectives: (i) 14 patches contained a high density of Brunner’s glands, which had previously been misclassified as crypts; (ii) 14 patches were selected due to poor performance of the prior segmentation model; and (iii) 12 patches were sourced from previously unused scanners to increase dataset heterogeneity and improve model robustness.

### Villus Morphology Training Dataset

The villus morphology dataset consisted of 41 WSIs scanned at 40× magnification (22 normal, 16 CD, 3 other diagnoses). A total of 3,354 villus manual annotations were generated in the form of three-point polylines (base, centre, and tip), created by four human annotators. These WSIs were collected from four hospitals and digitized using four different scanners. Age and sex information was available for 32 cases (78%), of which 22 (69%) were female, with a median age of 55 years.

### Classification Training and Validation Dataset

The classification training and validation dataset comprised 545 WSIs without pixel-level annotations. Each case was assigned a diagnosis based on the corresponding clinical pathology report. This dataset included 163 CD cases, 232 normal cases, and 150 cases with other non-coeliac pathological diagnoses. Data were collected from four hospitals and scanned using three different scanners. The median age was 60 years and 305 (57%) were female.

### Classification Test Dataset

The independent classification test dataset consisted of 1,357 WSIs, also without pixel-level annotations, with diagnoses derived from clinical pathology reports. This dataset included 250 CD cases, 795 normal cases, and 312 cases with other non-coeliac pathological diagnoses, including chronic inflammation, gastric metaplasia, and gastric heterotopia. Gastric metaplasia refers to replacement of the duodenal epithelium with gastric-type mucosa, while gastric heterotopia denotes ectopic gastric tissue within the duodenum. These conditions were included because they represent common non-coeliac findings in duodenal biopsies [28] that may share overlapping histological features with coeliac disease, including elevated IEL counts and villous architectural distortion [29]. Importantly, all cases originated from two hospitals not represented in the training datasets, and images were digitised using two scanners, including one from a manufacturer not present in the training data. Demographic information was available for all cases, with 56% female patients and a median age of 49 years.

## Methods

The proposed pipeline comprises five sequential stages as illustrated in Figure 1: (1) pre-processing, (2) semantic segmentation of villi, crypts, IELs, and enterocytes, (3) crypt instance segmentation and morphological feature extraction, (4) villus length estimation, and (5) coeliac disease diagnosis via a supervised classifier.

### 1 Pre-Processing

The majority of pre-processing steps follow our previous work [27]. WSIs are first processed using Schreiber’s method [30] to detect tissue regions and remove artefacts. The tissue mask is then used to extract non-overlapping patches of size 4096×4096 pixels at 40× magnification (approximately 0.25 µm per pixel). Patches are retained only if at least 25% of their area overlaps with the tissue mask. All retained patches are subsequently stain-normalised using the Macenko method [31].

### 2 Semantic Segmentation Model

As in our previous work [27], we employ two separate semantic segmentation models: one targeting larger tissue structures (villi and crypts), and a second focusing on cellular components (IELs and enterocytes). The IEL-enterocyte model is identical to that used in our prior study, whereas the villus-crypt model is retrained with an expanded training set, including an additional 40 manually annotated patches as described above.

For the villus–crypt model, we still use a U-Net++, but replace the ResNet-34 encoder with a ResNeSt-50d backbone, initialised with ImageNet-pretrained weights using the *timm* library. ResNeSt architectures incorporate a split-attention mechanism that has been shown to improve performance over standard ResNet architectures across a range of computer vision tasks [32].

Consistent with our previous work, the villus-crypt model is trained using 3-fold cross validation on patches of size 2048×2048 pixels, downsized to 20x magnification and with the same hyper-parameters as before. At inference time, however, we operate on larger 4096×4096 patches to improve spatial context, this is possible because U-Net++ architecture is agnostic to input size.

We additionally compare this approach to a pathology foundation model-based segmentation method using hibou-b [33]. However, this did not demonstrate superior performance compared to the U-Net++ ResNeSt-50d implementation, as detailed in Supplementary Material A.

Finally, we apply a post-processing procedure to refine the villus-crypt segmentation masks. This includes morphological operations to close small gaps and remove spurious components. Full implementation details and hyperparameters are provided in Supplementary Material B.

### 3 Crypt Morphological Analysis

We next aim to quantify crypt morphology by estimating the length, width, orientation, and eccentricity of individual crypts. This process consists of two stages: (i) conversion of the crypt semantic segmentation mask into an instance segmentation map, and (ii) extraction of morphological features for each detected crypt.

To obtain individual crypt instances, we derive an instance segmentation from the binary crypt mask produced by the semantic segmentation model. Due to variability in biopsy orientation, crypts may appear either approximately circular (cross-sectional view) or elongated/oval (longitudinal view), as visualised by Taavela et al. [9]. To account for this, we employ a two-branch algorithm that separately detects round and elongated crypts, followed by merging of the resulting instances.

For round crypts, we first fill internal holes within connected components of the binary mask to obtain solid regions. We then apply morphological erosion to separate adjacent structures, followed by a watershed segmentation to delineate individual crypt instances. Candidate regions are subsequently filtered based on a circularity criterion to retain approximately round structures.

For elongated crypts, we apply a sequence of alternating erosion and dilation operations to enhance separation between adjacent crypts while preserving elongated structures. This is followed by watershed segmentation to obtain individual instances. The results from both branches are then combined to produce the final instance segmentation mask.

For each detected crypt, we compute a set of morphological features, including centroid location (the geometric centre of the segmented crypt region), area, major and minor axis lengths (corresponding to crypt length and width), orientation, and eccentricity (a measure of deviation from circularity). These features are derived from the fitted ellipse of each instance.

A detailed description of the algorithm, including all implementation details and hyperparameters, is provided in Supplementary Material C.

### 4 Villus Morphological Analysis

We next aim to detect individual villi and quantify their morphology, in particular villus length. To achieve this, we propose a three-stage pipeline: (i) a polyline prediction model that localises keypoints along each villus, (ii) extraction of keypoints from predicted heatmaps, and (iii) matching of keypoints to form individual villus polylines.

#### 4.1. Polyline Model

The polyline model is designed to predict characteristic keypoints along each villus, namely the base, centre, and tip. The model takes as input 512 × 512 image patches at 5× magnification, with four input channels: RGB intensities and a single-channel villus–crypt semantic segmentation mask with discrete pixel values of 0, 1, and 2 corresponding to background, villi, and crypts, respectively.

The network outputs feature maps of size 128 × 128 (corresponding to 1.25× magnification) with four channels: three heatmaps encoding the likelihood of base, centre, and tip keypoints, and a single-channel embedding map used to group keypoints belonging to the same villus.

We adopt a CentreNet-based architecture [34], which is well suited to heatmap-based keypoint detection. The model is trained using a combination of heatmap regression loss and an embedding loss, that enforces intra-villus embedding consistency and inter-villus separation.

##### 4.1.1 Loss function

The total loss ℒ for each patch is defined as the sum of a heatmap loss ℒ_heat_ and an embedding loss ℒ_*embed*_:

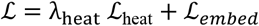

where λ_heat_ = 0.1 is the heatmap loss weight. The heatmap loss ℒ_heat_ is defined as the sum of losses over the three keypoint types (base, centre, tip):

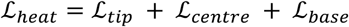

Each term is computed using a focal-style loss applied to Gaussian-smoothed ground truth heatmaps. For each *x* ∈ {tip, centre, base}:

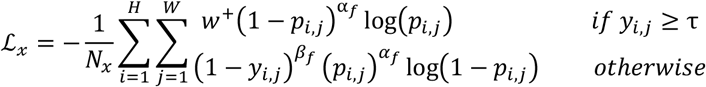

Where *H* = 128 *W* = 128 denote the height and width of the output heatmap, *y*_*i,j*_ ∈ [0,1] is the ground truth Gaussian-smoothed heatmap value at pixel (*i, j*) with parameter σ = 8, then *p*_*i,j*_ ∈ [0,1] is the predicted heatmap value at pixel (*i, j*). A_*f*_ = 2 is the focal loss exponent β_*f*_ = 4 down-weights near-peak background regions, τ = 0.95 is the positive sample threshold, *w*^+^ = 50 is a positive class weight to handle keypoint sparsity, *N*_*x*_ = ∑_*i,j*_ 𝟙[*y*_*i,j*_ ≥ τ] is the number of positive samples for keypoint type *x*.

The embedding loss consists of two components: a pull loss ℒ_pull_, which encourages embeddings of keypoints (base, centre, and tip) belonging to the same villus to be similar, and a push loss ℒ_push_, which encourages embeddings of different villi to be separated by at least a margin δ:

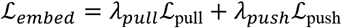

where *λ*_*pull*_ = 2 and *λ*_*push*_ = 1.

The pull loss is defined as:

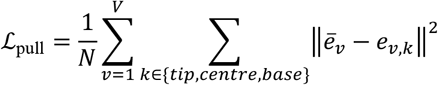

And the push loss is defined as:

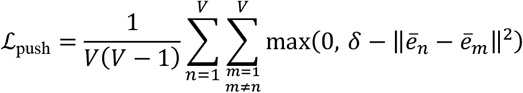

where *V* denotes the number of villi, and *N* is the total number of keypoint instances across all villi. The embedding vector at keypoint *K* of villus *𝒱* is denoted by *e*_v,k_, and the mean embedding for villus *𝒱* is given by:

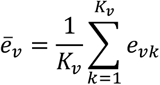

where *K*_v_ is the number of annotated keypoints for villus *𝒱*. The margin parameter is set to δ = 5, which enforces a minimum separation between embeddings of different villi.

Training was performed using three-fold cross-validation. Models were optimised with Adam (learning rate 8 × 10^-4^, weight decay 1 × 10^-4^) for 30 epochs with a constant learning rate and a batch size of 10. During training, random spatial augmentations were applied, including translations (≤5%), scaling (±10%), and rotations (≤15°), with constant padding. Horizontal and vertical flips were applied with probability 0.5.

#### 4.2. Keypoint extraction

Given the predicted heatmaps (normalised to lie within 0 and 1), we extract discrete keypoints corresponding to villus bases, midpoints, and tips. For each heatmap channel, activations below a fixed threshold (set to 0.4) are suppressed to reduce noise.

Connected component analysis is then applied independently to each thresholded heatmap to identify spatially contiguous regions of high activation. For each connected component, a single keypoint is selected as the pixel with the maximum heatmap response within that component. This results in a set of candidate base, midpoint, and tip villus keypoints for each patch (Figure 4).

**Figure 3.**
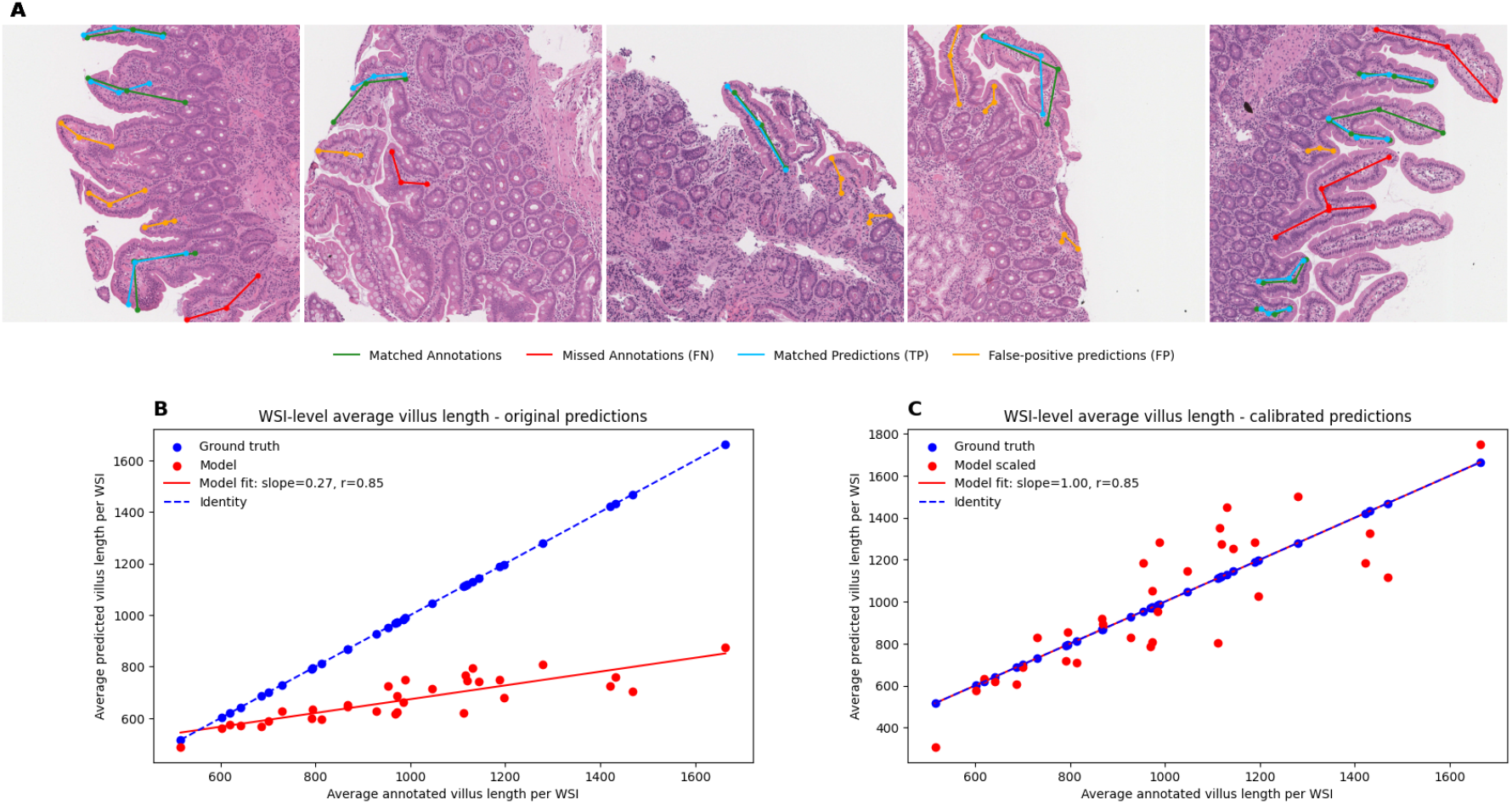
Villus detection performance and WSI-level length calibration. (A) Representative image patches showing matched detections, false negatives, and false positives. Green indicates matched annotations/predictions, red indicates missed villi, and orange indicates false-positive predictions. (B) Correlation between average villus length measured from manual annotations and raw model outputs at the WSI level. (C) Correlation after linear rescaling of model outputs.

**Figure 4.**
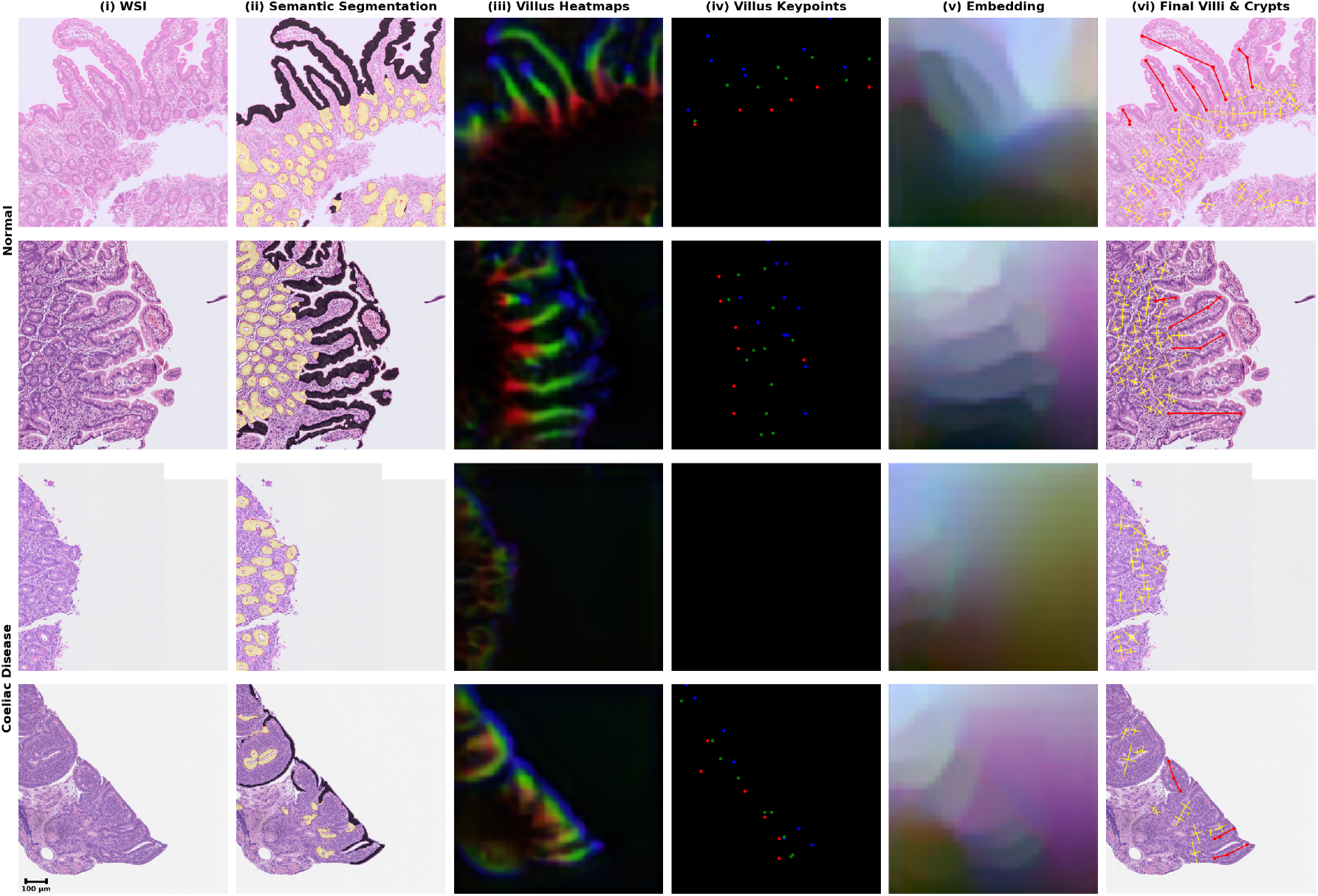
Visual examples of model outputs across normal and coeliac disease cases. The top two rows show normal biopsies, while the bottom two rows show coeliac disease biopsies. Columns show: (i) H&E WSI patches; (ii) semantic segmentation outputs highlighting villi (purple) and crypts (yellow); (iii) villus heatmaps produced by the polyline model, highlighting predicted villus tip (blue), centre (green), and base (red) regions; (iv) villus keypoints extracted from the heatmaps (v) embedding representations generated by the villus polyline model and used to associate detected keypoints belonging to the same villus instance; and (vi) final pipeline outputs showing detected villus polylines (red) and crypt major axes (yellow). Scale bar: 100 μm.

#### 4.3. Keypoint matching

We next associate extracted keypoints into base-midpoint-tip triplets, each corresponding to a single villus. To this end, we compute pairwise matching costs between base–midpoint and midpoint–tip pairs based on the embedding output of the polyline model. Specifically, the cost between two keypoints is defined as the absolute difference between their embedding values, averaged across the three cross-validation models. Candidate pairs with a cost exceeding a predefined threshold are discarded. Keypoints that are not part of any valid pair are subsequently pruned.

We then construct villus candidates by evaluating all feasible base–midpoint–tip triplets. Each triplet is assigned a total cost equal to the sum of its constituent pairwise costs. A greedy selection procedure is applied to identify a set of non-overlapping triplets with minimal total cost, where a large penalty is assigned to unmatched keypoints. Additionally, geometric constraints are enforced to prevent intersecting polylines.

We also explored a graph neural network-based matching approach; however, this yielded inferior performance compared to the proposed method (see Supplementary Material D).

### 5. Diagnostic Classifier

We develop a diagnostic classifier that predicts coeliac disease status (positive vs. negative) based on quantitative features derived from the preceding models. The classifier operates on three features computed at the WSI level.

First, the IEL-to-enterocyte ratio inside the villi is calculated from the semantic segmentation outputs and log-transformed to reduce skew. Second, the villus-to-crypt area ratio, also derived from semantic segmentation, is similarly log-transformed. Third, a villus-length-to-crypt-depth ratio is computed as the mean villus length divided by the mean crypt major axis length, capturing structural alterations in mucosal architecture.

These features are used as input to a logistic regression classifier with L2 regularisation (c=1) fitted using the L-BFGS solver (maximum of 1000 iterations). Model training is performed using a leave-one-source-out cross-validation scheme on the dataset that spans data from four different hospitals to assess generalisation across data from different acquisition sources. We select the value that optimises the mean of the true positive rate and the true negative rate as the decision threshold for predicting coeliac disease positive and coeliac disease negative outputs.

In cases where multiple WSIs are available for a single patient (e.g. biopsies obtained from both the duodenal bulb (D1) and the descending duodenum (D2)), predictions are first generated independently for each WSI. A patient-level diagnosis is then assigned as positive if at least one WSI is classified as coeliac disease positive.

We further evaluate the contribution of individual features through a comprehensive ablation study, including analysis of feature subsets, model configurations, and aggregation strategies (see Supplementary Material F). In addition, we quantify the discriminative power of each feature using Cohen’s d and statistical significance testing.

## Results

### 1. Semantic Segmentation Villus-Crypt Model

We trained semantic segmentation models using 3-fold cross validation to detect crypts and villi on 94 unique patches of size 1024×1024 at 20x magnification. The models achieved average validation performance of segmenting the background with 97% precision and 96% recall, villi with 87% precision and 89% recall, crypts with 88% precision and 90% recall.

On the 54 patches that were included in the previous study the new sets of models improved villus precision by 20% (67% to 87%) and recall by 9% (80% to 89%). For crypts precision improved by 10% (78% to 88%) and recall by 19% (71% to 90%).

Full confusion matrices for each individual fold as well as those averaged over all folds can be found in Supplementary Materials G. Examples outputs can be found in Figure 4.

### 2. Crypt Morphology

We developed a crypt morphology algorithm that takes as input the villus-crypt semantic segmentation mask and outputs, for each detected crypt, its length, width, orientation, and eccentricity. Visual examples of instance-level crypt segmentation and morphological feature extraction are provided in Figure 4, demonstrating reliable detection of individual crypts across a range of biopsy orientations and pathological diagnoses. The crypt major axis length is used to compute the villus-length-to-crypt-depth ratio in combination with the villus polyline model outputs. As shown in Figure 5, this ratio demonstrates strong discriminative ability between coeliac disease and all non-coeliac diagnostic groups across internal and external datasets, including normal mucosa and non-coeliac inflammatory conditions such as chronic inflammation, gastric metaplasia, and gastric heterotopia (p<10^−5^ in all comparisons on the test set).

**Figure 5.**
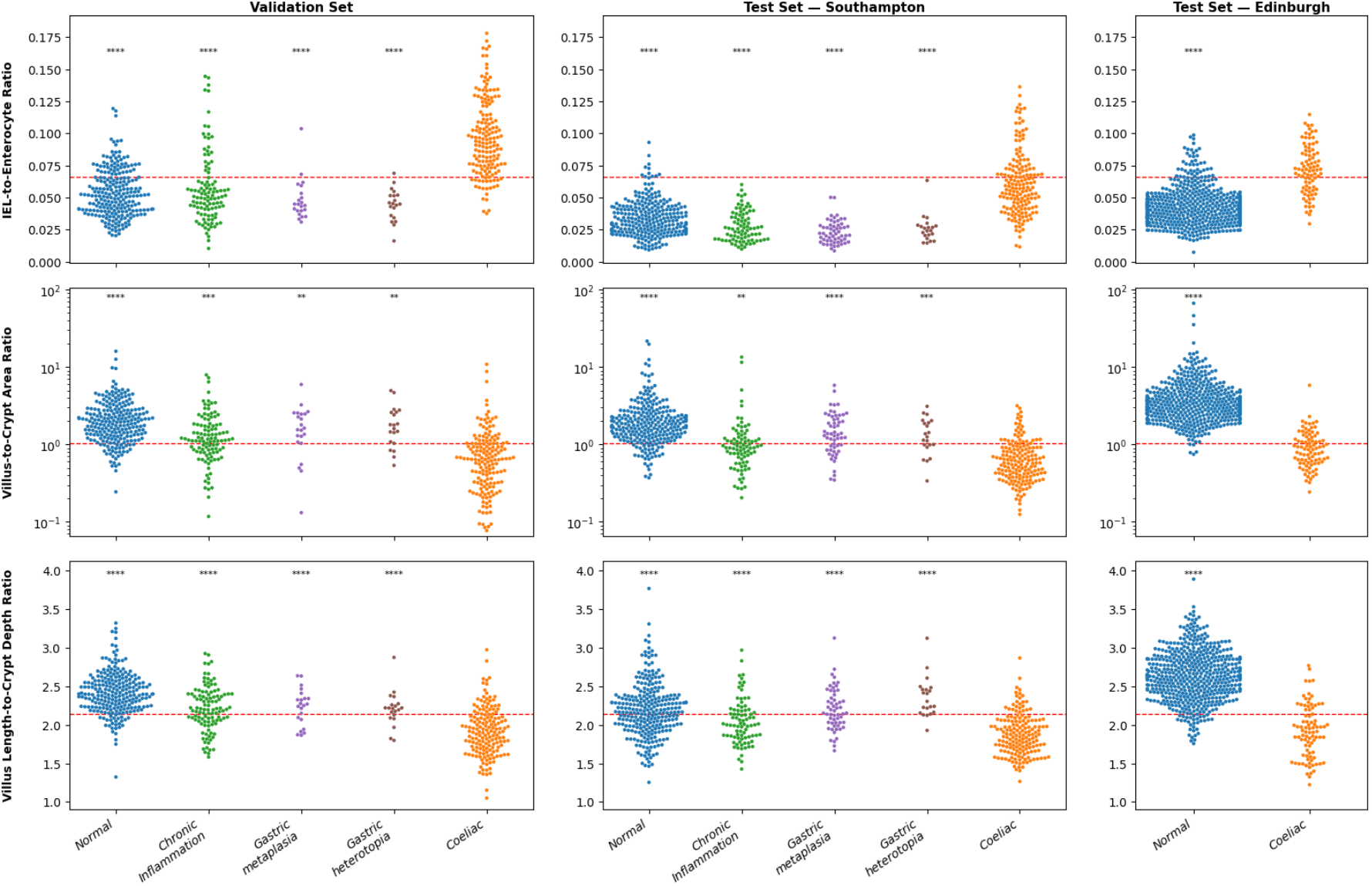
Swarm plots showing distributions of three WSI-derived features across diagnostic groups in the validation cohort (left), Southampton test cohort (middle), and Edinburgh external test cohort (right). Top row: IEL-to-enterocyte ratio. Middle row: villus-to-crypt ratio (log scale). Bottom row: villus length-to-crypt depth ratio. Each point represents a single WSI. Red dashed lines indicate the predefined diagnostic thresholds for each feature. Statistical comparisons were performed between coeliac disease and each non-coeliac diagnostic category using Welch’s t-test. Significance levels are denoted as: p < 0.05 (*), p < 0.01 (**), p < 0.001 (***), and p < 0.0001 (****).

### 3. Villus Morphology

#### 3.1 Keypoint Prediction

We trained polyline detection models to identify individual villi on a manually annotated dataset of 41 WSIs using three-fold cross-validation.

We first evaluated the initial stage of the villus polyline pipeline, which predicts three classes of villus keypoints: bases, centres, and tips. Each keypoint class was analysed independently. As predicted and annotated keypoints are unlikely to occur at exactly the same pixel location, predicted keypoints were matched to ground-truth annotations using Hungarian matching. A maximum matching distance of 80 μm was used, corresponding to approximately 320 pixels at 40x magnification and 10 pixels at the 1.25x magnification used for model inference. This threshold is small relative to a typical healthy villus length (500–1500 μm [35]), ensuring that only spatially proximate keypoint pairs are considered matches.

Matched predicted–annotated keypoint pairs were considered true positives. Predicted keypoints without a corresponding annotation were counted as false positives, while annotated keypoints without a matched prediction were counted as false negatives. Using this approach, we computed precision, recall for keypoint detection across all three keypoint classes. Overall precision was 40% and recall was 70%. The model was intentionally optimised towards higher recall at the expense of precision, as unmatched predicted keypoints are subsequently filtered during the villus triplet assembly stage.

For matched keypoint pairs, the mean localisation error was 29 μm. Villus tip predictions demonstrated the lowest localisation error (26 μm), followed by villus centres (32 μm) and villus bases (33 μm).

#### 3.2. Keypoint matching

We next evaluated how accurately predicted villus triplets corresponded to expert annotations. A predicted triplet was considered matched to a ground-truth triplet if any of the six constituent keypoints from either triplet lay within 200 pixels (50 μm) of a segment connecting two keypoints in the opposite triplet (e.g. the predicted centre point lying within 200 pixels of the annotated tip–centre segment). Examples of matched and unmatched annotated and predicted villi are shown in Figure 3a.

Using this matching criterion, we computed true positives, false positives, and false negatives at the villus level, obtaining a precision of 41.4%, and a recall of 73.9%. These metrics assess whether predicted and annotated villi correspond spatially and do not account for geometric similarity between matched structures.

We next quantified geometric agreement between matched villi. The mean absolute distance between matched predicted and annotated villi lengths was 64.1 μm, corresponding to a mean relative difference of 25.1%. At the patch level, comparing the mean villus length across all villi within a patch resulted in a mean absolute difference of 80.5 μm and a mean relative difference of 30.1%.

Finally, we assessed concordance between manually annotated and model-predicted average villus lengths at the whole-slide image level. Predicted measurements demonstrated strong correlation with expert annotations (Pearson’s r=0.85; Figure 3b), although the model systematically underestimated villus length relative to human annotations.

To correct for this calibration bias, we fitted a linear regression model between predicted and annotated average villus lengths and applied the inverse regression transform to rescale model predictions. Following calibration, the mean absolute error was reduced to 33.6 μm with a mean relative error of 13.5% (Figure 3c).

### 4. Feature Discriminability

We evaluated whether the outputs of the three models could distinguish coeliac disease from other duodenal conditions. Using the classification training and validation dataset across four hospitals, we compared the distributions of: (i) IEL-to-enterocyte ratio (IEL-enterocyte segmentation model), (ii) villus-to-crypt area ratio (villus-crypt segmentation model), and (iii) villus-length-to-crypt-depth ratio (villus polyline and crypt morphology models) across coeliac disease (n=163) and four non-coeliac diagnoses: normal (n=232), chronic or non-specific inflammation (n=108), gastric metaplasia (n=22), and gastric heterotopia (n=20); they are among the most common non-coeliac pathological findings in duodenal biopsies [28].

All three metrics showed significantly different distributions between coeliac disease and every other diagnostic group (Welch’s t-test, p<0.01 in all comparisons). Separation was particularly strong against normal tissue, with p<10^−41^, p<10^−17^, and p<10^−44^ for IEL-to-enterocyte ratio, villus-to-crypt area ratio, and villus-length-to-crypt-depth ratio respectively (Figure 5; full p-values in Supplementary Material E).

These findings generalised to both hospitals in the classification test dataset (Figure 5). On the Edinburgh test set, unseen during training or validation, all three metrics remained highly discriminative between normal (n=524) and coeliac disease (n=87), with p<10^−24^ across all comparisons. On the Southampton dataset, acquired on a different scanner manufacturer, coeliac disease (n=166) was again separable from all four non-coeliac groups (269 normal, 84 chronic or non-specific inflammation, 56 gastric metaplasia, 21 gastric heterotopia), with p<0.01 across all metrics and diagnostic groups. Notably, villus-length-to-crypt-depth ratio achieved stronger separation than villus-to-crypt area ratio on this dataset, suggesting it may be more robust to cross-scanner variability. Full p-values are reported in Supplementary Material E.

### 5. Diagnostic Model

We trained a binary logistic regression classifier with L2 regularisation on the classification training and validation dataset (545 WSIs; 163 CD, 232 normal, 150 other non-coeliac diagnoses including chronic inflammation, gastric metaplasia, and gastric heterotopia) using leave-one-source-out cross-validation to assess generalisation across acquisition sites. The models were designed to classify WSIs as either coeliac disease positive or negative using three quantitative features derived from the preceding models: (i) the IEL-to-enterocyte ratio obtained from the IEL-enterocyte segmentation model, (ii) the villus-to-crypt area ratio obtained from the villus-crypt segmentation model, and (iii) the villus-length-to-crypt-depth ratio derived from the villus polyline and crypt morphology models.

Evaluation was performed on an external test set collected from two hospitals not represented in the training cohort, including one site using a scanner manufacturer unseen during training. The model achieved an accuracy of 94.5%, positive predictive value (PPV) of 92.9%, negative predictive value (NPV) of 94.7%, and an area under the receiver operating characteristic curve (AUC) of 0.982. These results demonstrate strong generalisation across independent institutions and scanner platforms. Full performance metrics are provided in Supplementary Material H and Figure 6.

**Figure 6.**
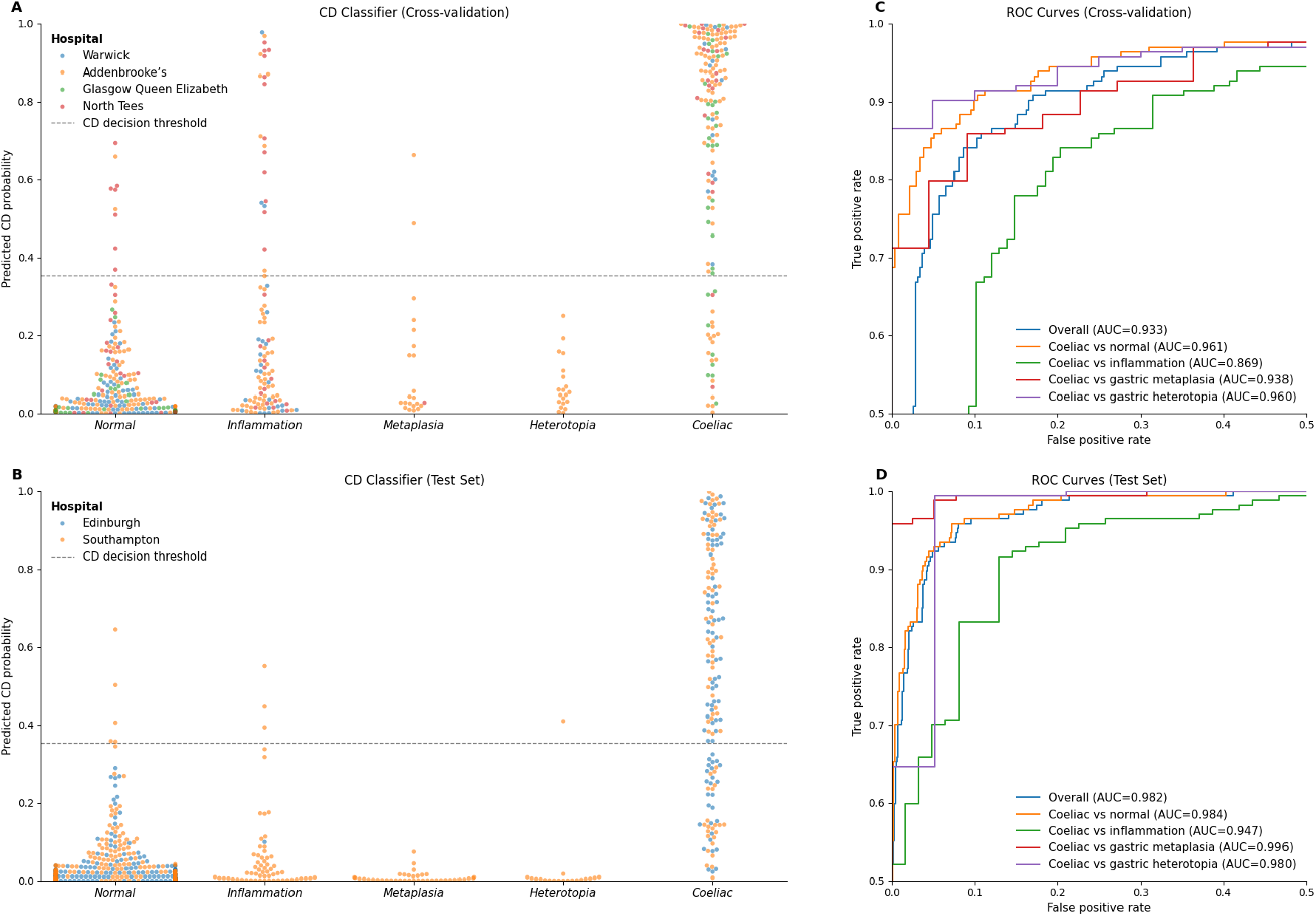
Diagnostic classifier performance across cross-validation and test datasets. (A) Predicted probabilities of coeliac disease generated by the diagnostic classifier for cases in the cross-validation cohort, stratified by diagnosis and hospital source. Each point represents a whole-slide image (WSI). The dashed horizontal line indicates the decision threshold used for classification, selected to maximise the average of the true positive rate and true negative rate. (B) Predicted probabilities for the test cohort, which includes hospitals not represented during training and Southampton cases acquired using a scanner manufacturer not included in the training data. (C) Receiver operating characteristic (ROC) curves for the cross-validation cohort, showing overall performance and pairwise comparisons between coeliac disease and individual non-coeliac diagnostic categories. (D) ROC curves for the external test cohort. The classifier achieved strong discrimination between coeliac disease and all non-coeliac categories, including chronic or non-specific inflammation, gastric metaplasia, and gastric heterotopia, across both internal cross-validation and external validation datasets.

Among the evaluated diagnostic comparisons, the lowest performance was observed when distinguishing coeliac disease from chronic inflammation, with an AUC of 94.7%. In contrast, comparisons between coeliac disease and normal mucosa, gastric metaplasia, or gastric heterotopia each achieved AUC values exceeding 98%.

Inference runtime and computational footprint analyses are reported in Supplementary Material I. The complete pipeline required approximately 240 seconds per whole-slide image on a single NVIDIA A100 GPU and was estimated to emit 0.0047 kgCO_2_ per diagnosed case. This represents less than 1% of the estimated carbon footprint associated with current diagnostic workflows.

## Discussion

In this study, we present an end-to-end interpretable deep learning framework for coeliac disease diagnosis from H&E-stained duodenal biopsies. Unlike most previous AI approaches for coeliac disease, which rely on black-box classification models [16–20], our framework explicitly extracts clinically meaningful morphological features including IEL-to-enterocyte ratios, villus morphology, and crypt morphology. These correspond to the key histopathological features routinely assessed by pathologists during coeliac disease diagnosis, enabling predictions that are transparent and clinically interpretable. Interpretability is increasingly recognised as essential for clinical adoption of AI systems in pathology and medical imaging more generally [21–23]. Existing explainability methods for black-box models, such as heatmaps, can provide some insight into model behaviour but often lack direct pathological interpretability. In contrast, our framework is inherently interpretable, as it produces pixel-level segmentation masks, villus polylines, and quantitative crypt measurements that directly reflect established histomorphological criteria used in routine pathology practice.

This is particularly relevant in coeliac disease, where histopathological assessment is known to have substantial inter-observer variability, with disagreement rates reported at approximately 20% [10–15]. By providing objective and quantitative measurements of villus length, crypt depth, and IEL frequency, the proposed framework may help improve diagnostic consistency and standardisation.

A major contribution of this work is the improved semantic segmentation model for villi and crypts. Compared with our previous study [27], the expanded training dataset and ResNeSt-50d backbone substantially improved segmentation performance and robustness across multiple centres and scanners: villus precision improved from 67% to 87% and recall from 80% to 89%, with corresponding crypt improvements from 78% to 88% precision and 71% to 90% recall. We additionally introduced a crypt morphology pipeline capable of extracting instance-level features including crypt length, width, orientation, and eccentricity, enabling direct quantification of crypt hyperplasia at the individual crypt level rather than relying solely on aggregate pixel-level measurements.

We further proposed a novel polyline-based model for detecting individual villi and estimating villus length, achieving strong WSI-level correlation with expert annotations (Pearson’s r=0.85) and a mean relative error of 13.5% after calibration. All three morphological features, IEL-to-enterocyte ratio, villus-to-crypt area ratio, and villus-length-to-crypt-depth ratio, showed highly significant separation between coeliac disease and all non-coeliac diagnostic groups across internal and external datasets (p<0.01 in all comparisons).

We developed a diagnostic classification model integrating IEL-to-enterocyte ratios, villus-to-crypt area ratios, and villus-length-to-crypt-depth ratios to predict coeliac disease status. On an independent external test set of 1,357 whole-slide images from two previously unseen institutions, including cases digitised with a scanner manufacturer not represented during training, the model achieved accuracy of 94.5%, PPV of 92.9%, NPV of 94.7%, and AUC of 0.982. This strong generalisation across institutions and scanner platforms compares favourably with the 82.66% accuracy reported by Tyagi et al. [26] for MeasureNet, which was not evaluated in a multi-centre setting. Importantly, strong performance was maintained across all five diagnostic categories including chronic inflammation, gastric metaplasia, and gastric heterotopia.

The main limitation of this work is that the ground truth diagnoses used for training and evaluation are derived from clinical pathology reports, which are themselves subject to the approximately 20% inter-observer disagreement inherent in coeliac disease histopathology, potentially introducing label noise. Additionally, while generalisation was demonstrated across multiple institutions and scanner manufacturers, further validation in routine clinical workflows would be required before deployment. Future work could address the challenge of branching villi, where a single villus base splits into multiple tips, which are not well represented by the current polyline formulation and likely contribute to the systematic underestimation of villus length relative to human annotations. In addition, the quantitative morphological features produced by the framework, such as crypt eccentricity and orientation, could be further explored to enable more direct assessment of biopsy orientation and tissue quality.

In conclusion, this work presents an interpretable AI framework for coeliac disease diagnosis that combines semantic segmentation, crypt morphometry, villus morphology estimation, and quantitative classification. The proposed system achieves strong performance across multi-centre datasets while providing clinically meaningful outputs aligned with established histopathological criteria. These results demonstrate a meaningful step toward transparent and reproducible AI-assisted diagnosis of coeliac disease.

## Data Availability

The patient data underlying this study are not publicly available.

## Acknowledgement

We would like to acknowledge Josephine Williams, Richard Walsh, Rosie Turnbull, Matthew Ellis, Singam Sivasankar, Michelle Watson, Avinash Thomas, and Obinna Enwereji for their support in collecting the data. We would further like to thank Graham Snudden for organisational support.

## Contributions

RB, MJA, EJS, and FJ conceived and designed the study. RB and JRD developed the AI model and performed the experiments. RB, AS, AS, GJ, and RR created annotations under the supervision of JYHC and MJA who provided expert gastrointestinal pathological input. FJ and RB drafted the manuscript and EJS, MJA, JYHC contributed to subsequent revisions and helped shape the final manuscript. All authors reviewed and approved the final version.

## HRA Approval

All slide scans and accompanying fully anonymised patient data were obtained with Health Research Authority (HRA) Approval (reference: 335851, issued on 18 December 2023) which stated that specific patient consent was not required for the study. The study was conducted in accordance with the principles of the Declaration of Helsinki and the UK Policy Framework for Health and Social Care Research.

## Data sharing statement

The patient data underlying this study are not publicly available.

