## Supplementary Materials for "Interpretable machine learning for coeliac disease diagnosis: quantitative morphometry of duodenal biopsies"

1. *Supplementary Material A – Semantic Segmentation Ablation* Study contains an ablation study comparing different backbone architectures for the villus-crypt semantic segmentation model.
2. *Supplementary Material B – Villus Crypt Semantic Segmentation Mask Refinement Algorithm* includes a detailed summary of the algorithm that refines the semantic segmentation mask returned by the Villus-Crypt Semantic Segmentation model.
3. *Supplementary Material C – Crypt Instance Segmentation Algorithm* details the algorithm that converts the semantic segmentation crypt mask into an instance segmentation mask.
4. *Supplementary Material D – Villus Polyline Model Ablation* Study includes an ablation study of the villus polyline model.
5. *Supplementary Material E – Feature Discriminability* highlights the ability for different features outputted by the various models to differentiate between normal and coeliac disease diagnoses.
6. *Supplementary Material F – Diagnostic Classifier Ablation Study* compares different architectures for the diagnostic classifier.
7. *Supplementary Material G – Villus-Crypt Semantic Segmentation Additional Results* contains more detailed results for the semantic segmentation model.
8. *Supplementary Material H – Diagnostic Classifier Additional Results* contains more detailed results of the diagnostic classifier.
9. *Supplementary Material I – Timings, Computational Requirements, and Carbon Footprint* includes a detailed calculation of the computational time requirements of our proposed software and the resulting carbon emissions of running our software in the UK.

### Supplementary Material A – Semantic Segmentation Ablation Study

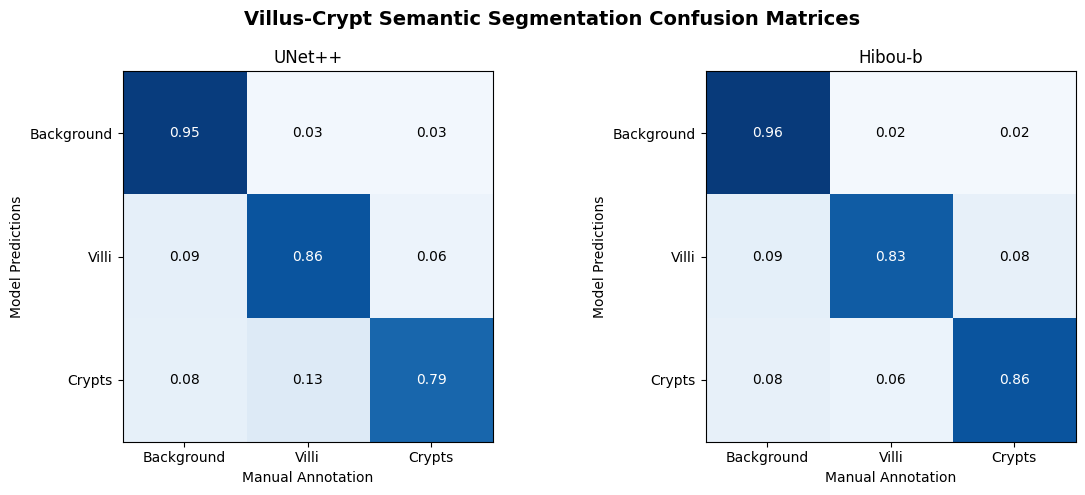

**Figure S1.** Confusion matrices comparing villus-crypt semantic segmentation performance between an ImageNet-pretrained U-Net++ model and the pathology foundation model Hibou-b. The Hibou-b backbone demonstrates improved semantic discrimination of crypt regions, particularly through increased crypt recall, reflecting stronger high-level tissue representations learned from pathology-specific pretraining.

| Feature | U-Net++ | Hibou-b |
| --- | --- | --- |
| IEL-to-Enterocyte area ratio | -1.763 | **-1.881** |
| Villus-to-Crypt area ratio | 1.337 | **1.313** |
| Villus length-to-crypt depth ratio | **1.224** | 0.858 |

**Table S1.** Comparison of feature discriminability between the U-Net++ and Hibou-b villus–crypt semantic segmentation models, quantified using Cohen’s *d* for separation of coeliac disease and normal cases (larger absolute values indicate stronger discriminability). The Hibou-b model shows modest improvements in pixel-derived features, including the IEL-to-enterocyte ratio and villus-to-crypt area ratio, but reduced performance for morphology-based villus length-to-crypt depth measurements.

As an additional experiment, we evaluated whether replacing the U-Net++ encoder with Hibou-b [1], a pathology-specific vision transformer foundation model, could improve semantic segmentation performance for villus–crypt analysis. Because Hibou-b is transformer-based rather than convolutional, several architectural modifications were required. Traditional U-Net architectures rely on skip connections to preserve spatial detail between encoder and decoder stages; to replicate this behaviour with a transformer encoder, we implemented a Dense Prediction Transformer (DPT)-style decoder. Hidden states from intermediate transformer layers (layers 3, 6, 9, and 12) were reshaped into multi-scale 2D feature maps and fused in a deep-to-shallow cascade with progressive upsampling, reintroducing fine-grained spatial information lost in deeper semantic representations. In addition, unlike U-Net++, Hibou was pretrained on fixed 224×224 image patches, requiring the segmentation framework to adopt a sliding-window inference strategy. Following empirical testing, 10x magnification patches were selected as the optimal compromise between contextual awareness and compatibility with the pretrained backbone.

The Hibou-based model achieves strong positive semantic segmentation results, particularly for crypt identification. Confusion matrices demonstrated improved recall of crypt regions compared with U-Net++, suggesting stronger high-level feature representations (Figure S1). Quantitative downstream analysis further showed modest improvements in pixel-level separation between normal and coeliac cases, particularly for villus-to-crypt and IEL-to-enterocyte area ratios. However, these gains did not consistently translate into improved geometric measurements of crypt morphology. Specifically, villus length-to-crypt depth separation was weaker for Hibou than for U-Net++ (Table S1).

Although Hibou produced semantically stronger predictions, the smaller 224×224 patch context reduced spatial awareness, leading to less geometrically coherent crypt boundaries. These segmentation irregularities propagated through the instance extraction and length measurement stages, negatively affecting morphometric accuracy. Consequently, despite the promising semantic improvements offered by the transformer backbone, U-Net++ was retained as the primary model in this work because accurate and robust crypt length extraction remained the central objective of the study.

### Supplementary Material B – Villus Crypt Semantic Segmentation Mask Refinement Algorithm

| **Algorithm S1: Crypt Semantic Segmentation Mask Refinement** | |
| --- | --- |
| 1 | **Input:** M ∈ {0,1,2}^{B×H×W} *▷ 0=background, 1=villus, 2=crypt* |
| 2 | **Output:** I, I_partial ∈ {0,1,2}^{B×H×W} *▷ fully & partially cleaned masks* |
| **Stage 1 — Morphological Pre-processing** | |
| 3 | *▷ Step 1: close small gaps caused by patch-boundary alignment errors* |
| 4 | M ← closing(M, disk(3)) |
| 5 | *▷ Save background mask for later restoration* |
| 6 | BG ← (M = 0) |
| 7 | *▷ Step 3: dilate crypt pixels to fill minor intra-crypt gaps, then close* |
| 8 | C_dil ← closing(dilation(M = 2, disk(7)), disk(7)) |
| 9 | *▷ Merge dilated crypts back with original villi* |
| 10 | M ← C_dil ∪ (M = 1) *▷ crypt pixels overwrite; villi fill the rest* |
| **Stage 2 — Fragment Removal & Partial Output (per image)** | |
| 11 | *▷ Step 2: discard all connected components smaller than 4,000 px* |
| 12 | M ← remove_small(M, min=4000 px) |
| 13 | *▷ Produce I_partial (saved before structure classification)* |
| 14 | I_partial ← M |
| 15 | I_partial[BG] ← 0 *▷ restore original background* |
| 16 | I_partial[I_partial = 2] ← remove_small( crypts, min=4000 px ) *▷ filter small crypt fragments* |
| **Stage 3 — Structure Labelling & Crypt Reclassification (per image)** | |
| 17 | *▷ Label every connected non-background component* |
| 18 | {R_i} ← label(M > 0) |
| 19 | **for each** structure R_i do |
| 20 | **if** area(R_i) ≥ 70,000 px **then** **skip** *▷ large structures need no reclassification* |
| 21 | *▷ Compute three classification criteria on bounding-box crop* |
| 22 | convex_hull ← convex_hull_image(R_i) |
| 23 | solidity ← area(R_i) / area(convex_hull) *▷ is structure convex?* |
| 24 | num_holes ← label(¬R_i) − 1 *▷ count enclosed holes* |
| 25 | euler ← 1 − num_holes *▷ euler < 1 → has a lumen* |
| 26 | n_crypt ← \|{ p ∈ R_i : M[p] = 2 }\| *▷ semantic crypt vote count* |
| 27 | *▷ All three criteria must pass to relabel as crypt* |
| 28 | **if** solidity > 0.3 **and** euler < 1 **and** n_crypt > 30 then |
| 29 | M[R_i] ← 2 *▷ relabel entire structure as crypt* |
| **Stage 4 — Post-processing (per image)** | |
| 30 | *▷ Restore original background* |
| 31 | M[BG] ← 0 |
| 32 | *▷ Discard small crypt fragments (second pass)* |
| 33 | M[M = 2] ← remove_small( crypts, min=4000 px ) |
| 34 | **return** M as I, I_partial |

The mask cleaning algorithm corrects common artefacts in the raw semantic segmentation output, including patch-boundary misalignments, intra-crypt gaps, and misclassified tissue regions, through a four-stage GPU-accelerated pipeline (Algorithm S1). First, morphological closing with a disk of radius 3 smooths patch-boundary discontinuities, the original background is saved for later restoration, and crypt pixels are dilated then closed with a disk of radius 7 to bridge minor intra-crypt gaps before being merged back with the original villus pixels. Second, connected components smaller than 4,000 pixels are discarded as noise fragments; a partially cleaned mask is saved at this point for downstream use before the more aggressive classification steps. Third, each remaining connected structure smaller than 70,000 pixels is evaluated against three criteria: a solidity greater than 0.3 (area relative to convex hull), an Euler number below 1 (indicating at least one enclosed hole consistent with a crypt lumen), and more than 30 crypt-labelled pixels from the original semantic prediction; structures satisfying all three are relabelled entirely as crypt, recovering instances that the semantic model had misclassified as villus. Finally, the saved background is restored and a second small-component removal pass filters any residual crypt fragments below 4,000 pixels, yielding the fully cleaned mask alongside the partially cleaned intermediate output.

### Supplementary Material C – Crypt Instance Segmentation Algorithm

We now present the crypt instance segmentation algorithm which converts a semantic segmentation mask into individually labelled crypt instances through two parallel pipelines that handle circular and elongated crypts separately (Algorithm S2). Circular crypts are isolated by filling luminal holes, eroding the mask to break touching boundaries, computing a normalised Euclidean distance transform thresholded at 0.35 to identify crypt centres as seed markers, and applying marker-controlled watershed to assign all crypt pixels to an individual instance; segmented regions are then filtered by their isoperimetric quotient, retaining only those with a roundness between 0.9 and 1.5. The remaining non-circular crypt pixels are processed by an iterative erosion scheme in which each connected component is eroded progressively until it splits into sub-components whose largest area falls below 1/1.7 of the original; when three or more sub-components emerge, a junction analysis dilates the two largest until they overlap and classifies the connecting bridge as either a thick shared wall, merging the pair into one crypt, or a thin junction separating two distinct crypts, based on the ratio of dilated component area to overlap area and the relative size difference between the two components. The resulting component labels are then used as markers for a second watershed pass to reconstruct full crypt bodies. Finally, the two instance masks are merged, crypts intersecting the image border are discarded, and ellipse-based region properties (centroid, area, axis lengths, orientation, eccentricity) are computed for each retained instance.

**Algorithm S2: Crypt Instance Segmentation**

**Input:** M ∈ {0,1,2}^{H×W} *▷ 0=background, 1=villus, 2=crypt*

**Output:** I ∈ ℤ≥0^{H×W} *▷ unique integer per crypt instance*

**Stage 1 — Pre-processing**

C ← (M = 2)	*▷ extract binary crypt mask*

C ← fill_holes(C)	*▷ close enclosed luminal holes*

**if** |C| = 0 **then return** ∅

**Stage 2 — Circular Crypt Segmentation**

*▷ Step 1: erode to separate touching boundaries*

E ← erode(C, k=3, n=5)

*▷ Step 2: distance transform → marker seeds*

D ← EDT(E)

D_norm ← (D − D_min) / (D_max − D_min)

𝒲 ← (D_norm > 0.35)	*▷ threshold: isolate crypt centres*

𝒮 ← label(𝒲)	*▷ connected-component markers*

*▷ Step 3: watershed — flood from seeds over −D, masked to C*

I_circ ← watershed(−D, 𝒮, mask=C)

*▷ Step 4: roundness filter ρ = P² / (4π A)*

**for each** region r in I_circ do

**if** 0.9 < P_r² / (4π A_r) < 1.5 **then** keep r	*▷ else remove*

| **Stage 3 — Long Crypt Segmentation** | |
| --- | --- |
| 18 | *▷ Step 1: mask out already-labelled circular crypts* |
| 19 | C_long ← C \ (I_circ > 0) |
| 20 | *▷ Step 2: erode to separate adjacent long crypts* |
| 21 | E_long ← erode(C_long, k=3, n=5) |
| 22 | *▷ Step 3: label connected components → candidate set {R_i}* |
| 23 | {R_i} ← label(E_long > 0) |
| 24 | *▷ Step 4 & 5: iterative erosion + junction analysis per candidate* |
| 25 | **for each** candidate R_i with area A_i do |
| 26 | **repeat** |
| 27 | R_i ← erode(R_i, k=3, n=4) *▷ aggressive erosion step* |
| 28 | sub ← connected_components(R_i) |
| 29 | **until** \|sub\| = 1 **or** area(R_i) < 1000 px **or** max_area(sub) < A_i / 1.7 |
| 30 | *▷ Junction analysis: triggered when ≥ 3 sub-components emerge* |
| 31 | **if** \|sub\| ≥ 3 **and** 2 smallest areas within 50% then |
| 32 | C1, C2 ← two largest sub-components |
| 33 | D1 ← dilate(C1, k=3, n=10) *▷ expand until they overlap* |
| 34 | D2 ← dilate(C2, k=3, n=10) |
| 35 | **while** D1 ∩ D2 = ∅ do |
| 36 | D1 ← dilate(D1, n=5) ; D2 ← dilate(D2, n=5) |
| 37 | A_J ← \|D1 ∩ D2\| *▷ overlap (junction) area* |
| 38 | thick_jn ← (\|D1\| < 3·A_J) or (\|D2\| < 3·A_J) |
| 39 | similar ← \|A_D1 − A_D2\| / max(A_D1,A_D2) ≤ 0.35 |
| 40 | **if** thick_jn and not similar then *▷ broad shared wall → one crypt* |
| 41 | label C1 ∪ C2 as single instance |
| 42 | **else** *▷ two distinct crypts* |
| 43 | label C1, C2 as separate instances |
| 44 | *▷ Step 6: watershed reconstruction over full long-crypt extent* |
| 45 | D_long ← EDT(𝒮_long > 0) |
| 46 | D_norm ← (D_long − D_min) / (D_max − D_min) |
| 47 | 𝒲_long ← (D_norm > 0.35) |
| 48 | I_long ← watershed(−𝒲_long, 𝒮_long, mask=C_long) |
| **Stage 4 — Combination & Post-processing** | |
| 49 | *▷ Step 1: merge (label indices are non-overlapping by construction)* |
| 50 | I ← I_circ + I_long |
| 51 | *▷ Step 2: remove border-touching crypts (within 2 px of image edge)* |
| 52 | ℬ ← { i : I[b] = i for some border pixel b } |
| 53 | I[ I ∈ ℬ ] ← 0 |
| 54 | *▷ Step 3: fit equivalent ellipse → region properties per instance* |
| 55 | 𝒫 ← regionprops(I, {centroid, area, major/minor axis, orientation, eccentricity}) |
| 56 | **return** I, 𝒫 |

*EDT = Euclidean Distance Transform. All morphological operations use a square structuring element of the stated kernel size.*

### Supplementary Material D – Villus Polyline Model Ablation Study

Several design decisions were explored during development of the villus keypoint detection model. First, three input configurations were compared: (i) only taking the RGB image as input, (ii) only using the villus-crypt semantic segmentation mask, (iii) and a combined 4-channel input combining both approaches. The 4-channel approach performed best. Second, two key parameters of the heatmap loss function were tuned: the positive sample threshold τ, which controls spatial specificity (set to 0.95 to keep predicted keypoints distinct without sacrificing generalisation), and the positive class weight w⁺ (set to 50 to counterbalance the dominance of background pixels). Third, Hungarian matching visualisations confirmed that recall consistently exceeds precision, which is expected: ground truth annotations only include fully visible villi, whereas the model correctly predicts keypoints for partially cropped villi at patch boundaries, producing additional predictions that are handled downstream by the triplet matching algorithm. Fourth, a preliminary experiment on the training set informed the pull-versus-push weighting in the loss function. And confirmed that learned embeddings improved triplet assignment accuracy compared to just using distance between points as the key metric. Finally, parameter sweeps over the keypoint and background thresholds of the triplet matching algorithm highlighted the inherent difficulty of selecting a single globally optimal threshold, as the best choice varied across images.

As an alternative to the hand-crafted triplet matching pipeline, we explored whether a graph neural network (GNN) could learn to link base, centre, and tip keypoints into villus triplets directly. Training a GNN on ground truth annotations alone is problematic, as ground truth contains no erroneous or unmatched keypoints, whereas the detection model over-predicts in practice. To address this, the keypoint detector was run on 36 training WSIs to obtain realistic keypoint sets, including spurious predictions, and these were then manually matched by an expert to produce a supervised training set with both matched and unmatched keypoints labelled.

The architecture is inspired by SuperGlue [2], a graph attention network (GAT) originally designed for cross-image point matching in 3D reconstruction. We adapted it by running two SuperGlue instances in parallel: one matching base-to-centre keypoints and one matching centre-to-tip keypoints. Final triplets are formed by chaining, retaining only (base, centre, tip) combinations where both a base to centre and a centre to tip match exist. Each keypoint was represented by its learned embedding (from the 3-fold detection model), spatial position, heatmap confidence value, and an edge bias initialisation. The edge bias provides the GNN with a spatial prior over which pairs of keypoints are likely to belong to the same villus: for each pair, the heatmap is binarised at four thresholds (0.1, 0.2, 0.3, 0.4) and the fraction of the line segment between the two points falling on heatmap-positive pixels is recorded, giving four coverage values that are compressed to a scalar via a small fully connected layer and added to the attention scores before softmax.

Performance was evaluated using Spearman ρ between per-WSI mean predicted villus length and per-WSI mean manually matched villus length across 32 WSIs, a rank-based metric chosen for its robustness to the systematic underprediction of absolute lengths caused by patch boundary truncation (longer villi are disproportionately cut off at patch edges, biasing predicted lengths downward relative to full-WSI ground truth). The best GNN achieved ρ = 0.749, compared to ρ = 0.772 for the original triplet matching pipeline. While the GNN did not surpass the existing pipeline within the time available for optimisation, we believe a well-optimised GNN remains a promising direction for future work.

### Supplementary Material E – Feature Discriminability

|  | Validation | Test Edinburgh | Test Southampton |
| --- | --- | --- | --- |
| IEL-to-enterocyte ratio | -1.83 | -1.86 | -1.45 |
| Villus-to-crypt-area ratio | 0.88 | 0.78 | 0.87 |
| Villus length-to-crypt-depth ratio | 1.74 | 2.19 | 1.01 |
| Villus length - mean | 0.99 | 1.5 | 0.7 |
| Villus length – 25^th^ quantile | 0.55 | 0.77 | 0.03 |
| Villus length – median | 0.81 | 1.21 | 0.41 |
| Villus length – 75^th^ quantile | 1.0 | 1.36 | 0.75 |
| Crypt majority axis length - mean | -1.37 | -2.1 | -0.79 |
| Crypt majority axis length – 25^th^ quantile | -0.82 | -1.5 | 0.1 |
| Crypt majority axis length - median | -1.28 | -2.26 | -0.67 |
| Crypt majority axis length – 75^th^ quantile | -1.2 | -1.87 | -0.89 |
| Crypt minority axis length – mean | -1.28 | -2.14 | -0.5 |
| Crypt eccentricity mean | 0.26 | 0.0 | -0.25 |
| Crypt Area | -1.45 | -2.39 | -0.94 |

**Table S2.** Cohen's *d* effect sizes for each morphological feature, computed between normal and coeliac cases across the validation set and both held-out test sets (Edinburgh and Southampton). Positive values indicate the feature is larger in normal tissue; negative values indicate it is larger in coeliac tissue. Conventional thresholds: |*d*| < 0.5 small, 0.5–0.8 medium, > 0.8 large.

Feature discriminability was assessed by computing Cohen's *d* between normal and coeliac cases for each morphological feature across all three datasets (Table S2). The IEL-to-enterocyte ratio was consistently the most strongly discriminating feature in the negative direction (*d* ≈ −1.5 to −1.9), reflecting the well-established elevation of intraepithelial lymphocytes in coeliac disease. Villus-related features showed large positive effect sizes across all datasets: the villus length-to-crypt-depth ratio (*d* = 1.01–2.19) and villus length statistics (*d* = 0.41–1.5) were among the strongest positive discriminators, consistent with the villus blunting and crypt hyperplasia that characterise coeliac pathology. Crypt size features, including major axis length, minor axis length, and crypt area, exhibited consistently large negative effect sizes (*d* ≈ −0.8 to −2.4), indicating that crypts take up a larger fraction of the entire biopsy in coeliac tissue. Crypt eccentricity showed weaker and less consistent discrimination across datasets (*d* = −0.25 to 0.26), which is expected as this is mostly a result of the biopsy orientation, rather than diagnosis. Effect sizes were broadly consistent between the validation set and both test sets, supporting the generalisability of these features across different patient cohorts and acquisition sites.

### Supplementary Material F – Diagnostic Classifier Ablation Study

| Classifier | Accuracy | Sensitivity | Specificity | PPV | NPV | AUC |
| --- | --- | --- | --- | --- | --- | --- |
| Logistic Regression | **0.909 ± 0.071** | **0.916 ± 0.074** | 0.913 ± 0.118 | 0.908 ± 0.126 | **0.919 ± 0.073** | **0.973 ± 0.021** |
| Random Forest | 0.903 ± 0.071 | 0.893 ± 0.101 | 0.925 ± 0.124 | **0.919 ± 0.132** | 0.903 ± 0.092 | 0.960 ± 0.028 |
| SVM | 0.892 ± 0.062 | 0.866 ± 0.110 | **0.932 ± 0.106** | 0.899 ± 0.117 | 0.884 ± 0.098 | 0.953 ± 0.037 |
| XGBoost | 0.874 ± 0.077 | 0.911 ± 0.091 | 0.858 ± 0.140 | 0.837 ± 0.121 | 0.912 ± 0.089 | 0.940 ± 0.047 |

*Table S3. Comparison of diagnostic classifier architectures on the Normal vs. Coeliac validation dataset (mean ± SD across cross-validation folds). Thresholds were optimised by F1 score in all cases.*

To evaluate the choice of diagnostic classifier, we compared four model architectures on the Normal vs. Coeliac validation dataset, with all decision thresholds optimised by F1 score. Logistic regression was trained with L2 regularisation (C=1) with feature scaling applied beforehand. The random forest comprised 150 trees with a maximum depth of 3 and a minimum of 5 samples per split, requiring no feature scaling given its tree-based nature. The SVM used an RBF kernel (C=10, gamma='auto') with balanced class weights and feature scaling. XGBoost was used with default gradient boosting parameters. Results are summarised in Table S3.

Logistic regression achieved the highest discriminability (AUC 0.973 ± 0.021), outperforming random forest (0.960 ± 0.028), SVM (0.953 ± 0.037), and XGBoost (0.940 ± 0.047), while also yielding the highest accuracy (0.909 ± 0.071). That the simplest and most interpretable model performs best likely reflects the well-characterised nature of coeliac disease: the key diagnostic features are relatively few and well-defined, such that a linear decision boundary is sufficient to separate the classes without the need for more complex modelling. Logistic regression was therefore adopted for all final results.

### Supplementary Material G – Villus-Crypt Semantic Segmentation Additional Results

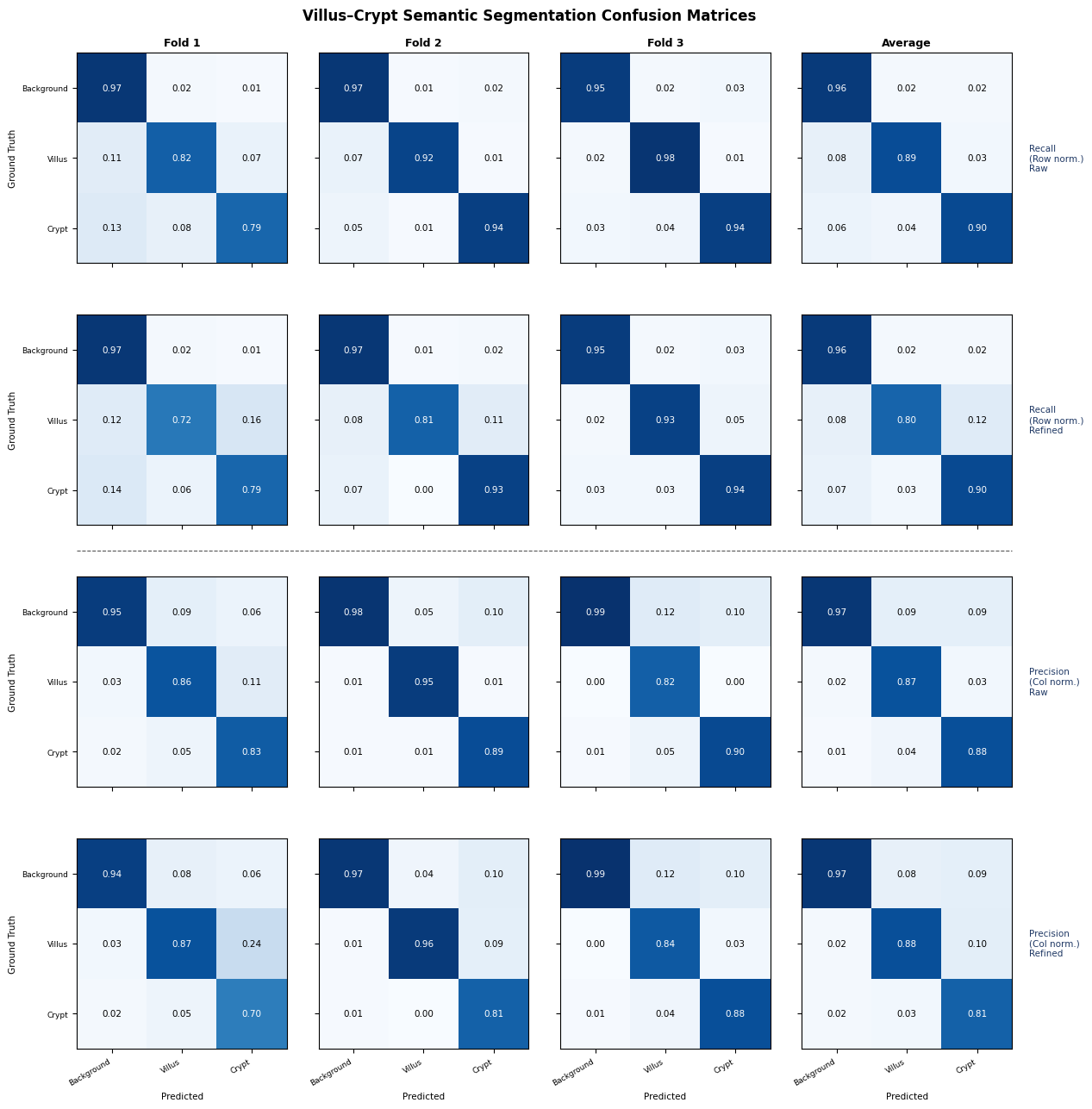

**Figure S2.** Confusion matrices for the villus-crypt semantic segmentation model evaluated under 3-fold cross-validation. Columns 1–3 show individual folds; column 4 shows the average across folds. Rows 1 and 2 are row-normalised (recall); rows 3 and 4 are column-normalised (precision). Odd rows show the raw model output; even rows show performance after application of the mask refinement algorithm.

Full per-fold and average confusion matrices for the villus-crypt semantic segmentation model are shown in Figure S2. Results are presented both before and after application of the mask refinement algorithm (Supplementary Material B), and under both row and column normalisation, corresponding to precision and recall respectively.

Across all folds, performance was consistent, with the averaged matrices (rightmost column) showing background precision and recall of 0.96-0.97, villus precision and recall of 0.80-0.89, and crypt precision and recall of 0.81-0.90. The primary source of confusion was between villi and crypts, with a small proportion of crypt pixels misclassified as villus and vice versa; background classification was highly stable across all folds. The refinement algorithm led to small increases in villus precision, but decreases in performance for crypt precision and villus recall.

### Supplementary Material H – Diagnostic Classifier Additional Results

|  | Accuracy | Sensitivity | Specificity | PPV | NPV | AUC |
| --- | --- | --- | --- | --- | --- | --- |
| Overall (n=1069) | 94.5% | 70.1% | 99.0% | 92.9% | 94.7% | 0.982 |
| Southampton (n=457) | 93.0% | 71.3% | 97.6% | 86.4% | 94.1% | 0.968 |
| Edinburgh (n=612) | 95.6% | 69.0% | 100% | 100% | 95.1% | 0.993 |
| CD vs normal (n=949) | 94.2% | 70.1% | 99.4% | 95.9% | 94.0% | 0.984 |
| CD vs chronic inflammation (n=229) | 76.9% | 70.1% | 95.2% | 97.5% | 54.1% | 0.947 |
| CD vs gastric metaplasia (n=206) | 75.7% | 70.1% | 100% | 100% | 43.8% | 0.996 |
| CD vs gastric heterotopia (n=186) | 72.6% | 70.1% | 94.7% | 99.2% | 26.5% | 0.98 |

**Table S4.** Additional diagnostic classifier performance metrics stratified by acquisition site and diagnostic subgroup. Metrics are reported for the external test cohort and include accuracy, sensitivity, specificity, positive predictive value (PPV), negative predictive value (NPV), and area under the receiver operating characteristic curve (AUC). Subgroup analyses demonstrate strong generalisation across institutions and scanner platforms, with highest performance observed for distinguishing coeliac disease from normal mucosa and gastric metaplasia, while chronic inflammation represented the most challenging differential diagnosis.

To further assess diagnostic robustness, classifier performance was stratified by hospital source and diagnostic subgroup (Supplementary Table S4). Performance remained consistent across both external institutions, including the Southampton cohort acquired using a scanner manufacturer not represented in the training data, demonstrating strong cross-site generalisation. The classifier achieved particularly high discriminative performance when distinguishing coeliac disease from normal mucosa, gastric metaplasia, and gastric heterotopia, with AUC values of 0.984, 0.996, and 0.980 respectively. The most challenging comparison was between coeliac disease and chronic inflammation (AUC 0.947), reflecting the greater morphological overlap between these entities. Across all subgroup analyses, specificity and positive predictive value remained high, indicating that false-positive predictions were uncommon and that the quantitative histological features extracted by the upstream segmentation and morphology models provided robust diagnostic separation across a range of clinically relevant differential diagnoses.

### Supplementary Material I – Timings, Computational Requirements, and Carbon Footprint

|  | Mean (s) | Standard deviation (s) | Min (s) | Max (s) |
| --- | --- | --- | --- | --- |
| Total image processing time | 239.99 | 153.07 | 5.29 | 843.68 |
| Tissue finder and patch extraction |  |  |  |  |
| IEL-enterocyte segmentation | 49.81 | 23.72 | 0.74 | 150.94 |
| Villus-crypt segmentation | 37.94 | 18.00 | 0.66 | 112.05 |
| Crypt refinement | 7.25 | 12.92 | 0.05 | 196.18 |
| Crypt instance segmentation | 33.05 | 18.14 | 0.12 | 109.22 |
| Villus Morphology | 1.84 | 1.08 | 0.03 | 8.41 |

**Table S5.** Computational runtime breakdown for the full diagnostic pipeline across the validation cohort. Reported values show the mean, standard deviation, minimum, and maximum execution time for each processing stage, including tissue detection, semantic segmentation, crypt refinement, instance segmentation, and villus morphology extraction. Timings were measured using a single NVIDIA A100-SXM-80GB GPU.

To estimate the computational footprint of the proposed pipeline, we measured inference times for each stage of processing across the validation cohort (Supplementary Table S5). The complete workflow required a mean processing time of 239.99 seconds per whole-slide image on a single NVIDIA A100-SXM-80GB GPU, corresponding to approximately 15 cases processed per hour. The IEL-enterocyte and villus-crypt semantic segmentation stages represented the largest computational components of the pipeline, while downstream morphology extraction and refinement steps contributed comparatively little to total runtime. Using the UK average electricity carbon intensity of 0.180 kgCO₂/kWh [3], the NVIDIA A100-SXM-80GB GPU emits approximately 0.07 kgCO₂ emissions per hour of operation [4]. The pipeline therefore produces an estimated 0.0047 kgCO₂ per diagnosed case. Extrapolated nationally, assuming approximately 400,000 duodenal biopsies are processed annually in the United Kingdom [5,6], full deployment of the algorithm would correspond to an estimated yearly carbon footprint of approximately 1,866 kgCO₂. These findings demonstrate that the computational requirements of the pipeline remain modest relative to large-scale clinical deployment.

Although direct comparison with human diagnostic workflows is difficult, published lifecycle analyses indicate that conventional pathology services are energy intensive due to continuous laboratory ventilation, refrigeration, staining workflows, and consumable usage, with routine histopathology processing estimated to emit approximately 0.6 kgCO₂e per slide [7]. In comparison, the proposed AI pipeline emits an estimated 0.0047 kgCO₂e per case during inference, suggesting that computational deployment contributes only a very modest (0.8%) additional environmental burden relative to standard pathology operations.
